# AI-based histologic scoring enables automated and reproducible assessment of enrollment criteria and endpoints in NASH clinical trials

**DOI:** 10.1101/2023.04.20.23288534

**Authors:** Janani S. Iyer, Harsha Pokkalla, Charles Biddle-Snead, Oscar Carrasco-Zevallos, Mary Lin, Zahil Shanis, Quang Le, Dinkar Juyal, Maryam Pouryahya, Aryan Pedawi, Sara Hoffman, Hunter Elliott, Kenneth Leidal, Robert P. Myers, Chuhan Chung, Andrew N. Billin, Timothy R. Watkins, Murray Resnick, Katy Wack, Jon Glickman, Alastair D. Burt, Rohit Loomba, Arun J. Sanyal, Michael C. Montalto, Andrew H. Beck, Amaro Taylor-Weiner, Ilan Wapinski

**Author notes:** **Corresponding author contact information:** Ilan Wapinski. Affiliation shown is that during the time of study; current affiliation is Johnson & Johnson, New Brunswick, NJ, USA. Affiliation shown is that during the time of study; current affiliation is AstraZeneca, Gaithersburg, MD, USA. Affiliation shown is that during the time of study; current affiliation is Atomwise, San Francisco, CA, USA. Affiliation shown is that during the time of study; current affiliation is BigHat Biosciences, San Mateo, CA, USA. Affiliation shown is that during the time of study; current affiliation is Genesis Therapeutics, Burlingame, CA, USA. Affiliation shown is that during the time of study; current affiliation is OrsoBio, Inc., Palo Alto, CA, USA. Affiliation shown is that during the time of study; current affiliation is Inipharm, San Diego, CA, USA. Affiliation shown is that during the time of study; current affiliation is Rhode Island Hospital and The Miriam Hospital, Providence, RI, USA.

## Abstract

Clinical trials in nonalcoholic steatohepatitis (NASH) require histologic scoring for assessment of inclusion criteria and endpoints. However, guidelines for scoring key features have led to variability in interpretation, impacting clinical trial outcomes. We developed an artificial intelligence (AI)-based measurement (AIM) tool for scoring NASH histology (AIM-NASH). AIM-NASH predictions for NASH Clinical Research Network (CRN) grades of necroinflammation and stages of fibrosis aligned with expert consensus scores and were reproducible. Continuous scores produced by AIM-NASH for key histological features of NASH correlated with mean pathologist scores and with noninvasive biomarkers and strongly predicted patient outcomes. In a retrospective analysis of the ATLAS trial, previously unmet pathological endpoints were met when scored by the AIM-NASH algorithm alone. Overall, these results suggest that AIM-NASH may assist pathologists in histologic review of NASH clinical trials, reducing inter-rater variability on trial outcomes and offering a more sensitive and reproducible measure of patient therapeutic response.

---

Nonalcoholic steatohepatitis (NASH), the progressive form of nonalcoholic fatty liver disease (NAFLD), is a rapidly growing cause of cirrhosis and hepatocellular carcinoma and is the most common indication for liver transplantation in women and older adults in the United States^1^. Despite the increasing incidence of NASH, including a ∼106% increase of cirrhosis caused by NASH from 1990 to 2017^2^, and the resulting medical and economic burden^3^, the development of effective therapeutics has proved challenging, and no effective pharmacologic treatment currently exists.

Histologically assessed endpoints are currently accepted as candidate surrogate endpoints in clinical trials evaluating therapeutics for NASH, and histologic criteria that indicate disease activity or severity are used as the basis for trial enrollment, risk stratification, and endpoint assessment in many different clinical indications. However, key aspects of trial design can be impacted by dependency on manual pathologist scoring of histologic features and limited by the sensitivity of scoring systems. Variability in assessment of histology-based endpoints and treatment-associated improvements can contribute to incomplete measurement of treatment response^4,5^, clinical trial failure^6^, difficulty in identifying an appropriate study population, and mistaken inclusion or exclusion of study participants^6–10^. Such errors could affect observed treatment responses and trial safety; for example, inappropriately included study participants may not respond to therapies or may respond more poorly than the intended population.

The U.S. Food and Drug Administration (FDA) and the European Medicines Agency (EMA) have issued guidance on the use of histopathologic assessment of liver biopsies as clinical trial inclusion criteria and change in histology-based scores over time as endpoints to measure trial outcomes to support accelerated approval of therapies for NASH^11–14^. Most histologic scoring systems proposed to date, with the NASH Clinical Research Network (CRN) used by the majority of studies and accepted by both the FDA and EMA, recommend measurement of four key features: macrovesicular steatosis, lobular inflammation, hepatocellular ballooning, and fibrosis^15–17^. Despite ongoing efforts by expert NASH pathologists to harmonize scoring guidelines^18^, both within clinical trial settings in their assigned panels and outside of specific trials as a community^7,16,18,19^, a recent re-analysis of a NASH clinical trial reported that a significant portion of the study population (23–31%) included in the trial did not meet enrollment criteria upon re-evaluation by a second hepatopathologist^6^. A separate investigation into the interpretation of hepatocellular ballooning by experienced pathologists showed high variation in the number of ballooned cells identified and no consensus on images that were free of ballooned hepatocytes^7^. This lack of reliability can reduce the power of NASH trials to detect a significant drug effect, as trials are not typically designed and powered to adequately account for variability in manual histologic scoring among pathologists.

Advances in artificial intelligence (AI) have led to the development of algorithms that can enable accurate, quantitative, and reproducible assessment of digitized pathology whole-slide images (WSIs)^5,20^. However, algorithms for the reliable detection, grading, and staging of NASH are not yet employed in clinical settings and have not received regulatory approval for clinical trial use. Here, we report a robust approach to evaluate assessment of NASH disease severity and improve clinical trial reliability by using a digital pathology tool for AI-based measurement (AIM) of NASH histology (AIM-NASH).

## Results

### Model-based evaluation of NASH histology

Machine learning (ML) models for segmenting and grading/staging NASH histologic features were trained using 103,579 pathologist-provided annotations of 8747 hematoxylin and eosin (H&E) and 7660 Masson’s trichrome (MT) WSIs from six completed phase 2b and phase 3 NASH clinical trials (Supplementary Table 1)^21–24^. The model development dataset described above was split into training (∼70%), validation (∼15%), and held-out test (∼15%) sets. The dataset was split at the patient level, with all WSIs from the same patient allocated to the same development set. Sets were also balanced for key NASH disease severity metrics, such as NASH CRN steatosis grade, ballooning grade, lobular inflammation grade, and fibrosis stage, to the greatest extent possible. The held-out test set contained a dataset from an independent clinical trial to ensure algorithm performance was meeting acceptance criteria on a completely held-out patient cohort in an independent clinical trial and to avoid any test data leakage^25^.

For each WSI, these models produced categories of histologic readouts generated separately by convolutional neural networks (CNNs) and graph neural networks (GNNs). AIM-NASH consisted of a pipeline in which CNNs were used to generate tissue overlays containing colorized predictions of segmentation indicating each histologic feature, and slide-level quantifications of the proportionate area of each feature. GNNs were used to predict a NASH CRN ordinal grade or stage for each histologic feature and a corresponding continuous score^26^ (Fig. 1 and Supplementary Fig.1). The overall process for the segmentation model development is shown in Supplementary Fig. 2.

**Fig. 1.**
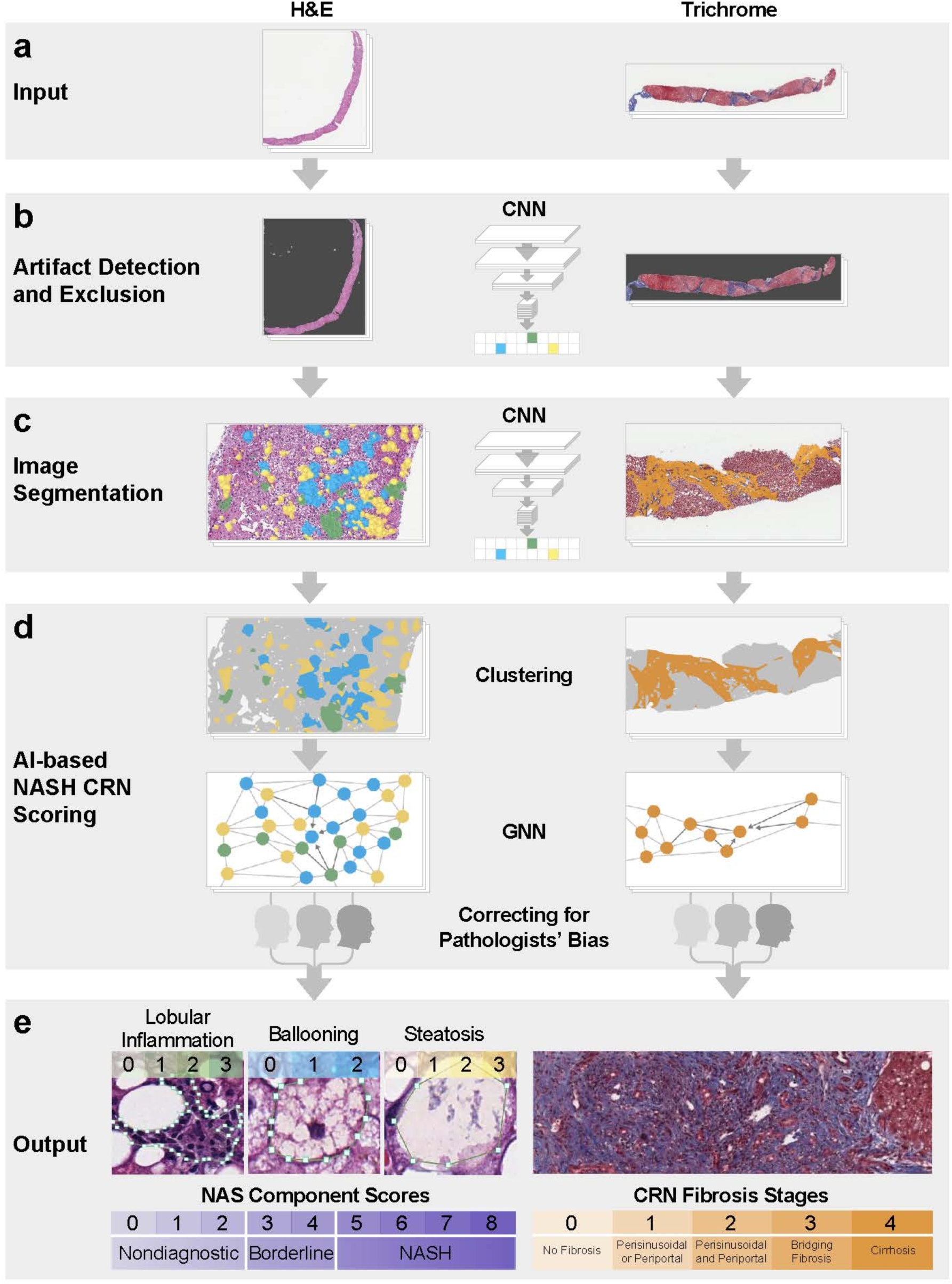
Pipeline for model deployment. (**a**) The model was trained with digitized H&E- and MT-stained images. (**b**) An artifact model detected image and tissue artifacts and excluded them prior to downstream analysis for both H&E and MT WSIs. (**c**) Pixel-level predictions of relevant histologic features were generated using a CNN trained on pathologist annotations. (**d**) Pixel-level predictions were clustered using GNNs. To correct for pathologists’ bias, the GNN model was specified as a “mixed effects” model, biases are learned, and GNNs are deployed with predictions using only the unbiased estimate. GNN nodes and edges were built from CNN predictions of relevant histologic features in the first model training stage. (**e**) This two-stage ML approach produced patient-level predictions of NASH CRN NAS component scores and fibrosis stage. AI, artificial intelligence; CNN, convolutional neural network; CRN, Clinical Research Network; GNN, graph neural network; H&E, hematoxylin and eosin; MT, Masson’s trichrome; NAS, nonalcoholic fatty liver disease activity score; NASH, nonalcoholic steatohepatitis; WSI, whole side image.

### Model outputs

#### Tissue overlays

CNNs were trained on annotations provided by 59 board-certified pathologists, with a subspeciality in liver pathology, as model inputs to identify key histologic features of interest on H&E and MT WSIs (Fig. 1a). An artifact model learned to distinguish evaluable liver tissue from features to be excluded from downstream analysis, including artifacts (e.g. tissue folds, out-of-focus areas) and WSI background (Fig. 1b). H&E CNNs segmented NAFLD Activity Score (NAS) component features (macrovesicular steatosis, hepatocellular ballooning, and lobular inflammation) and other relevant features, including portal inflammation, microvesicular steatosis, interface hepatitis, and normal hepatocytes (not exhibiting steatosis or ballooning). MT CNNs were trained to segment large intrahepatic septal and subcapsular regions (nonpathologic fibrosis), pathologic fibrosis, and bile ducts (Fig. 1c). Model-derived predictions for location and extent of each artifact and tissue feature per slide were displayed as colorized overlays over the original WSIs, facilitating quality control of the model’s prediction accuracy for each histologic feature (Fig. 2).

**Fig. 2.**
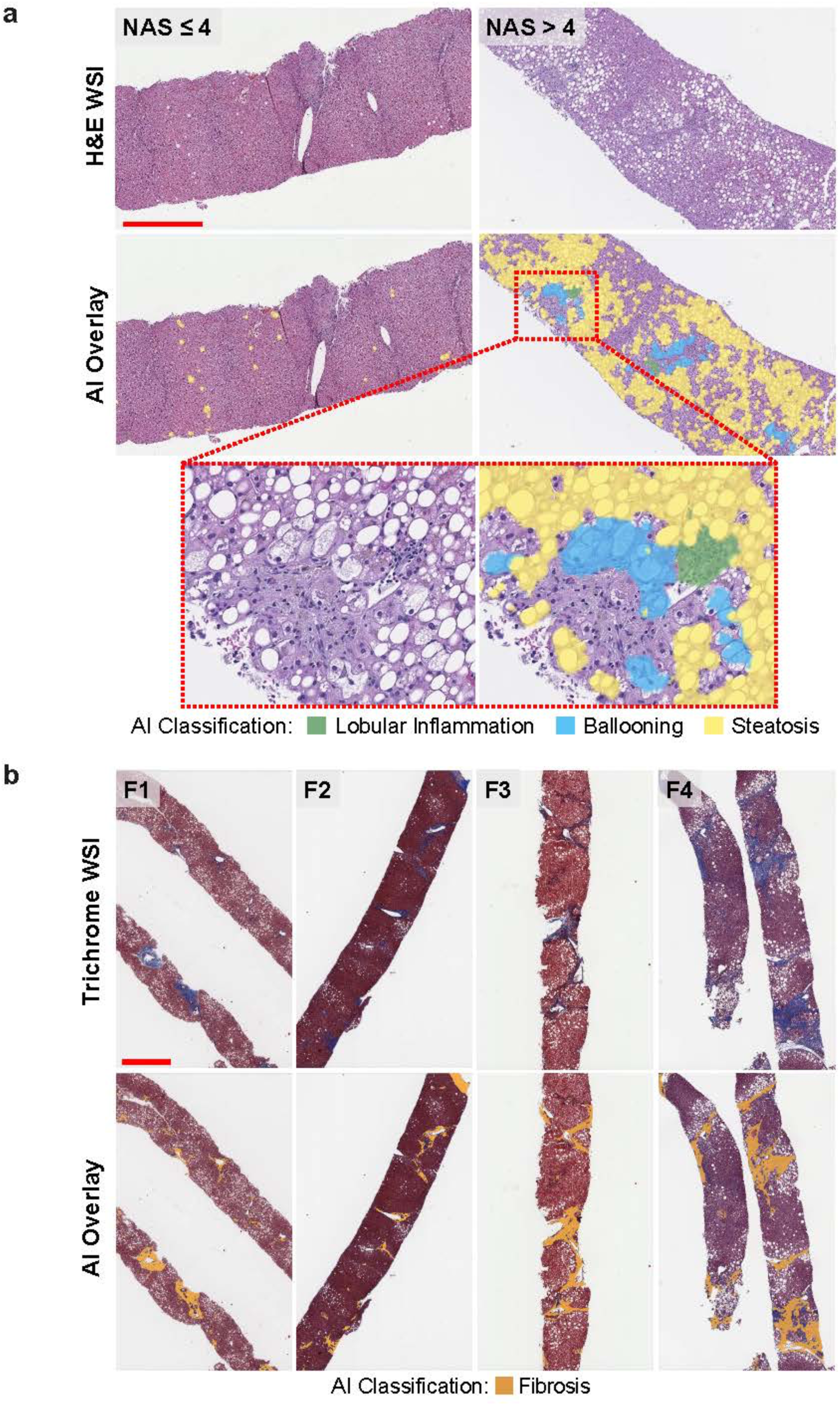
AI-based detection and scoring of NAS components and fibrosis. The NASH algorithm can detect histopathologic features on WSIs across a range of NASH disease severity. (**a**) Representative H&E-stained slides show AI overlays highlighting regions of steatosis, lobular inflammation, and ballooning. Cases corresponding to NAS ≤ 4 and NASH > 4, according to both pathologist consensus scoring and AI, are shown. The inset is a magnified field showing the presence of the three NAS components. (**b**) Representative MT-stained slides of each NASH CRN fibrosis stage show AI-generated overlays highlighting regions of fibrosis present on biopsies. Cases corresponding to NASH CRN fibrosis stages F1–F4, according to both pathologist consensus scoring and AI, are shown. These AI-generated overlays allow for qualitative review of model performance. Scale bar is 1 mm. AI, artificial intelligence; CRN, Clinical Research Network; H&E, hematoxylin and eosin; F, fibrosis stage; MT, Masson’s trichrome; NAS, nonalcoholic fatty liver disease activity score; NASH, nonalcoholic steatohepatitis; WSI, whole slide image.

#### Histologic feature proportionate area measurements

CNN-derived histologic feature predictions were quantified to generate slide-level area measurements per feature. These measurements were expressed both as raw area quantities (mm^2^) and as percentages of relevant histology and artifact normalized relative to total usable artifact-free tissue area in the WSI. Artifact-proportionate area measurements enabled efficient slide-level quality assessments and exclusion of inadequate image areas (e.g. artifacts, out of focus areas, tissue folds). Proportionate area measurements for H&E and MT NASH features, such as steatosis, ballooning, inflammation, and fibrosis, were used to evaluate disease severity.

#### Model-derived predictions for NASH CRN ordinal and continuous grades and stages

Spatially resolved overlays from CNN image segmentation algorithms were used as inputs, and pathologists provided slide-level NASH CRN grades/stages as labels to train GNNs (see Online Methods). GNNs were trained to predict NASH CRN steatosis grade, lobular inflammation grade, and hepatocellular ballooning grade from H&E-stained WSIs, and fibrosis stage from MT-stained WSIs (Fig. 1d,e). To generate interpretable, high-resolution NASH CRN grades and stages, GNN-predicted scores calculated on a continuum were mapped to bins, each equivalent to one grade or stage (Supplementary Fig. 3). For example, the continuous range for steatosis 0–1 was mapped to ordinal CRN grade 0, 1–2 to CRN grade 1, 2–3 to CRN grade 2, and 3–4 to CRN grade 3.

### Model performance repeatability and accuracy

In initial model performance testing relevant for application to both enrollment criteria and endpoints, the AIM-NASH algorithm scoring was highly reproducible. For each of the four histologic features, a comparison of 10 independent AIM-NASH reads per WSI resulted in a model versus model agreement rate of 100% (κ = 1; Supplementary Table 2), in contrast to modest intra-pathologist agreement using conventional approaches^6^. AIM-NASH performance accuracy was assessed using the mixed leave-one-out (MLOO) approach (see Online Methods). Comparing model performance with a pathologist-based consensus for each of the four histologic features, model versus consensus agreement rates fell within the range of previously reported rates of inter-pathologist agreement (Table 1)^16,19^. The model versus consensus agreement rate was greatest for steatosis (κ = 0.74, 95% confidence interval [CI] 0.71–0.77), followed by ballooning (κ = 0.70, 95% CI 0.66–0.73), lobular inflammation (κ = 0.67, 95% CI 0.64–0.71), and fibrosis (κ = 0.62, 95% CI 0.58–0.65). In addition, agreement between the model and consensus was greater than agreement for any individual pathologist against the other three reads, and greater than any mean pairwise pathologist agreement (Table 1).

**Table 1.** Model performance accuracy assessment. AIM-NASH performance was tested on an external, held-out dataset comprising WSIs from a phase 2b NASH clinical trial. Agreement rates for AIM-NASH grades/stages versus a consensus of three expert pathologists were superior to mean agreement between any individual pathologist and a panel comprising the other two pathologists and the model, and superior to any mean pairwise pathologist agreement. AIM, artificial intelligence-based measurement; CI, confidence interval; NASH, nonalcoholic steatohepatitis; WSI, whole side image.

| Agreement rate (95% CI) |  |  |  |
| --- | --- | --- | --- |
| Histologic feature | AIM-NASH versus consensus | Mean pathologist versus consensus | Mean pairwise pathologist agreement |
| Lobular inflammation | 0.67 (0.64–0.71) | 0.64 (0.62–0.67) | 0.58 (0.55–0.6) |
| Ballooning | 0.70 (0.66–0.73) | 0.66 (0.63–0.69) | 0.61 (0.59–0.64) |
| Steatosis | 0.74 (0.71–0.77) | 0.69 (0.66–0.72) | 0.62 (0.6–0.65) |
| Fibrosis | 0.62 (0.58–0.65) | 0.59 (0.57–0.62) | 0.54 (0.51–0.56) |

AIM, artificial intelligence-based measurement; CI, confidence interval; NASH, nonalcoholic steatohepatitis; WSI, whole slide image.

### Clinical utility of model-derived histologic grading and staging

#### AI-based evaluation of clinical trial enrollment criteria and endpoints

For patients with noncirrhotic NASH and fibrosis, the FDA has proposed criteria for NASH clinical trial enrollment and endpoint assessment^13^. For phase 2b and 3 trials, the FDA recommends criteria such as histologic diagnosis of NASH and fibrosis made within 6 months of enrollment, NAS ≥ 4 with inflammation and ballooning, CRN fibrosis score > 1 and < 4, among others. Recommended NASH clinical trial endpoints include evidence of efficacy using a histologic endpoint (late phase 2b trials) and resolution of NASH, fibrosis, or both NASH and fibrosis for phase 3 trials.

To demonstrate clinical relevance, AIM-NASH was deployed on WSIs from a completed phase 2b NASH clinical trial^22^ to generate scores based on histologic criteria and to identify patients eligible for enrollment (Supplementary Table 1, Analytic performance test set). AI-derived predictions for each of the patient cohorts were compared with the same cohorts identified by the clinical trial’s central readers (*n* = 3).

Model-derived histologic predictions were used to calculate NASH CRN scores and distinguished NAS ≥ 4 (with each component grade ≥ 1) from NAS < 4, criteria used to determine trial enrollment (Supplementary Table 1). The AIM-NASH versus consensus percent agreement (0.82; 95% CI 0.79– 0.85) was comparable with that of an average pathologist versus consensus (0.81; 95% CI 0.78–0.83; Fig. 3a). A similar result was observed for fibrosis. For distinguishing fibrosis stages F1–F3 versus F4, the model versus consensus agreement was 0.97 (95% CI 0.95–0.98), similar to the average pathologist versus consensus agreement of 0.96 (95% CI 0.95–0.97; Fig. 3a).

**Fig. 3.**
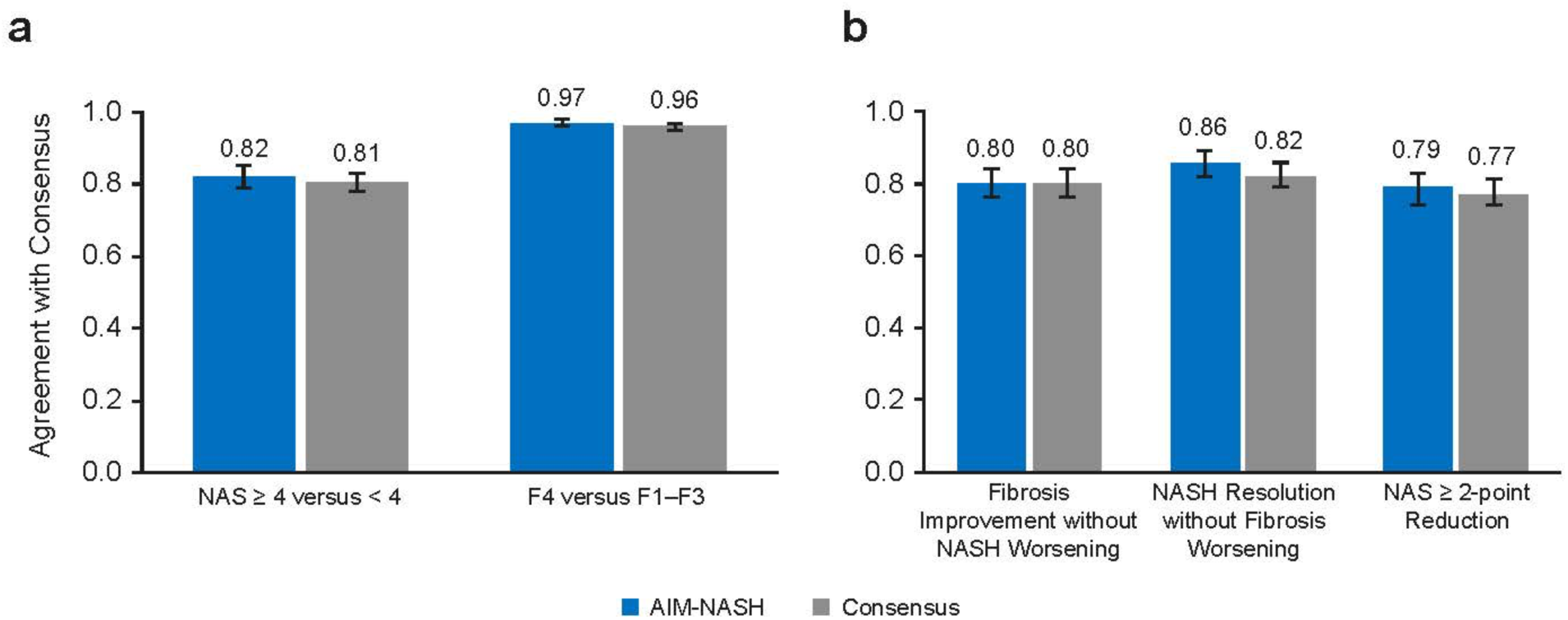
AI-based grading/staging of enrollment criteria and efficacy endpoints. (**a**) Model-derived scores distinguished fibrosis stages F1–F3 versus F4 and NAS ≥ 4 (with each component grade ≥ 1) versus NAS < 4, criteria used to determine trial enrollment. AIM-NASH agreement with consensus was comparable with that of each pathologist. (**b**) For assessment of efficacy endpoints commonly used in phase 2b and phase 3 NASH clinical trials (including NASH resolution without worsening of fibrosis, fibrosis improvement without worsening of NASH, and improvement in the composite NAS), AIM-NASH agreement with consensus was comparable with that of an average pathologist. Assessment was performed on an external held-out validation dataset from a phase 2b NASH clinical trial. AI, artificial intelligence; AIM, artificial intelligence-based measurement; F, fibrosis stage; NAS, nonalcoholic fatty liver disease activity score; NASH, nonalcoholic steatohepatitis.

Next, AIM-NASH was used on an enrolled NASH trial dataset to determine component scores and evaluate composite endpoints in a retrospective, exploratory manner. As part of the retrospective analysis of WSIs from the phase 2b NASH clinical trial^22^, histologic changes from baseline measured by AIM-NASH were compared with a consensus determination of the endpoints by three expert pathologists. Overall, AIM-NASH-based grading and staging for histologic endpoint assessment were comparable with those of mean individual pathologist versus consensus (Fig. 3b). For fibrosis improvement without worsening of NASH, both AIM-NASH versus consensus and pathologist versus consensus percent agreement rates were 0.80 (model versus consensus 95% CI 0.76–0.84; pathologist versus consensus 95% CI 0.77–0.83). For NASH resolution without worsening of fibrosis, the model versus consensus agreement rate (0.86, 95% CI 0.82–0.89) was moderately greater than the pathologist versus consensus agreement (0.82, 95% CI 0.79–0.86). A similar result was observed for a ≥ 2-point reduction in NAS, where the model versus consensus agreement rate (0.79, 95% CI 0.74–0.83) was comparable to the pathologist versus consensus agreement rate (0.77, 95% CI 0.74–0.81).

#### AI-based detection of treatment response in NASH clinical trials

To demonstrate AIM-NASH’s ability to measure treatment response in NASH clinical trials, we performed a retrospective analysis of drug efficacy in the ATLAS phase 2b clinical trial,^27^ which evaluated the combination of cilofexor (CILO) and firsocostat (FIR) in monotherapy and combination (CILO+FIR) in patients with advanced (F3–F4) fibrosis, leveraging the study’s original primary (≥ 1-stage improvement in fibrosis without worsening of NASH between baseline and week 48, as assessed by a single central pathologist [CP]) and exploratory endpoints. Although no treatment arm achieved statistical significance for the primary endpoint, the cohort that received the combination of CILO+FIR showed the greatest improvement in histology relative to placebo^27^.

To assess whether AIM-NASH could also detect histologic score–based improvement in patients who received the CILO+FIR treatment in the ATLAS trial, we deployed the AIM-NASH models on digitized WSIs from baseline and week 48 biopsies from enrolled patients. Model predictions for ordinal NASH CRN grades/stages were generated and compared with CP grades/stages for the trial endpoints. In addition to computing the proportion of responders per endpoint, treatment arm, and evaluation method, we also computed the difference in proportion of responders between CILO+FIR and placebo (placebo-adjusted response rate).

This analysis showed that compared with the CP, AIM-NASH detected a greater proportion of treatment responders in the CILO+FIR group for all three endpoints measured (≥ 1-stage fibrosis improvement without NASH worsening, 27% versus 19%; NASH resolution without fibrosis worsening, 24% versus 5%; ≥ 2-point reduction in NAS, 60% versus 35%; Fig. 4a), in addition to showing a numerically greater response in treated patients relative to placebo for all three endpoints (Fig. 4b).

**Fig. 4:**
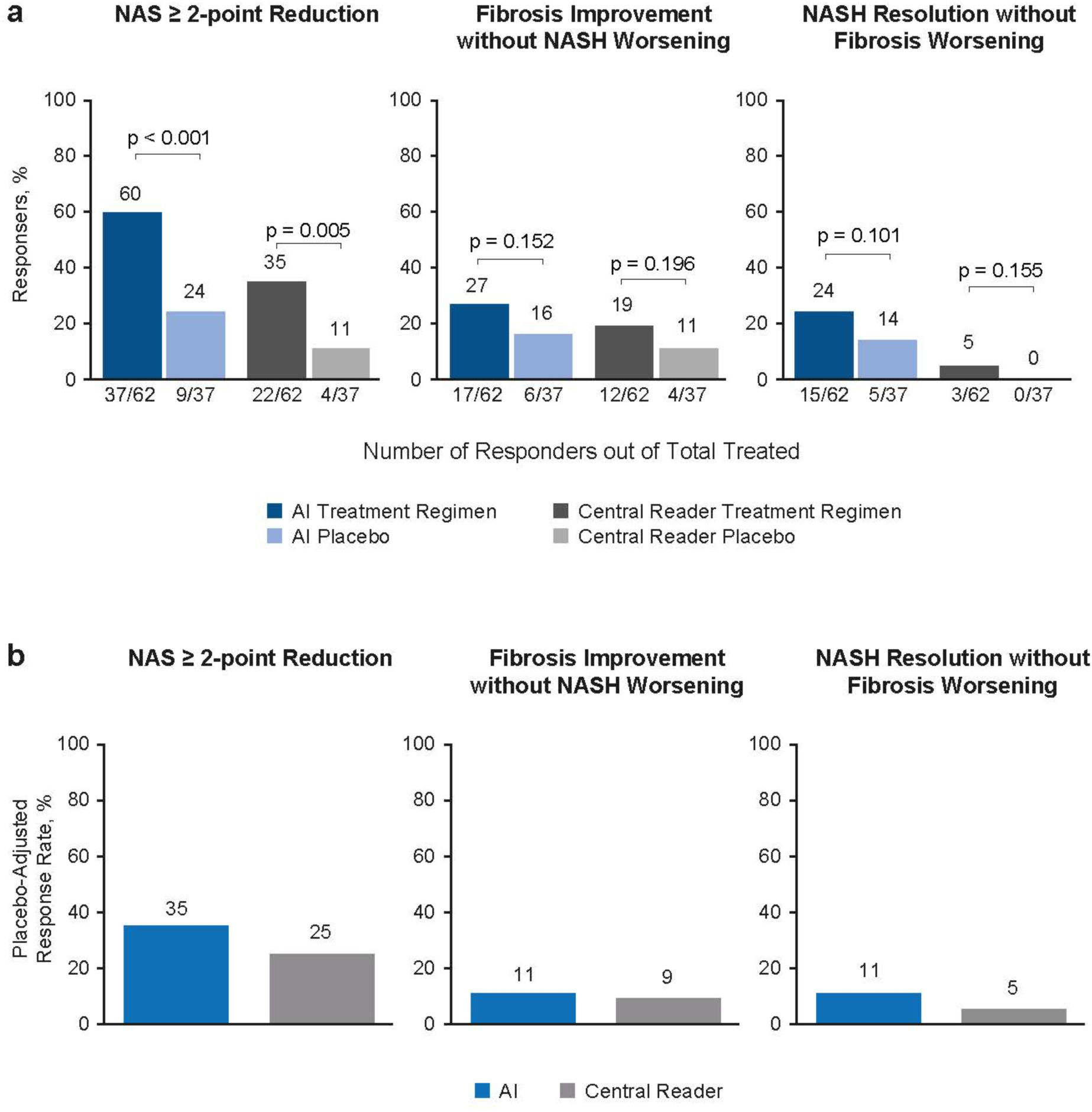
AIM-based retrospective drug efficacy assessment. AIM-NASH models were deployed on WSIs from baseline and week 48 biopsies from patients enrolled in the phase 2b ATLAS trial, which evaluated combination therapies for individuals with advanced NASH fibrosis. (**a**) For the trial endpoint NAS ≥ 2-point improvement, fibrosis improvement without worsening of NASH, and NASH resolution without worsening of fibrosis, AIM-NASH models showed a greater proportion of responders compared with that determined by the trial central reader. Sample sizes varied depending on data availability. (**b**) The placebo-adjusted response rate detected by AIM-NASH was greater than that detected by the central reader. AI, artificial intelligence; AIM, artificial intelligence-based measurement; NAS, nonalcoholic fatty liver disease activity score; NASH, nonalcoholic steatohepatitis; WSI, whole slide image.

### AI-enabled continuous scoring of NASH CRN components

We developed a continuous scoring system that enabled sensitive detection of changes in histology on a continuous scale and was mapped directly to the ordinal system, facilitating interpretation and seamless navigation between the ordinal and continuous systems when assessing therapeutic effect in NASH clinical trials (Supplementary Fig. 3).

#### Biological relevance of continuous scoring

AI-enabled continuous scoring was evaluated by correlating continuous scores against mean scores from three pathologists in a held-out validation dataset. We observed that continuous scores were significantly correlated with mean pathologist scores, confirming alignment between ML-derived continuous scores and directional bias of panel-based pathologist scoring (Fig. 5a). These results suggest that the disease severity indicated by the underlying tissue biology in each WSI was similarly captured through subordinal measurements, both by AIM-NASH and by the panel of pathologists, but could not be captured by a single pathologist providing ordinal scores for staging and grading scores (Fig. 5a).

**Fig. 5.**
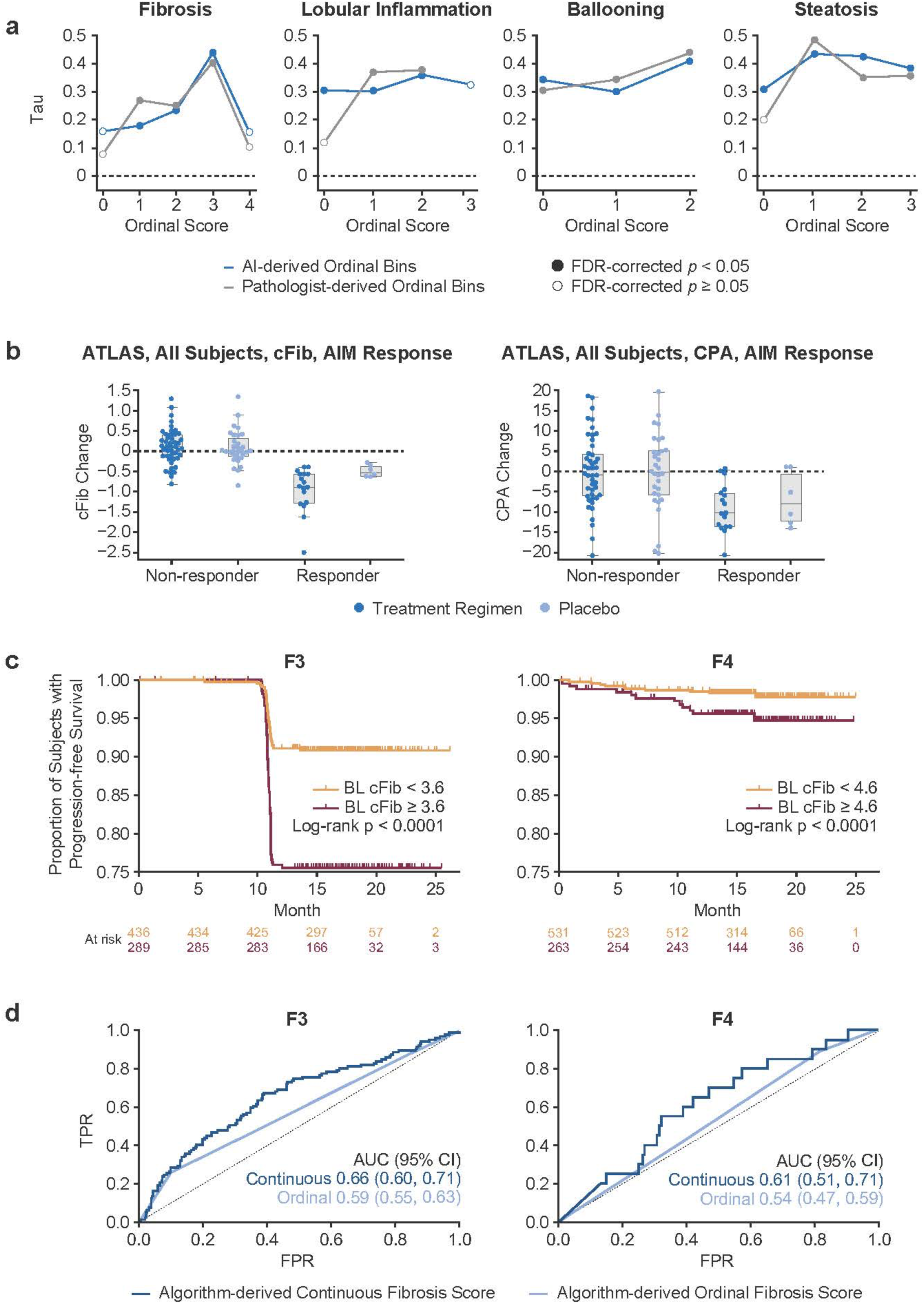
AI-based continuous NASH CRN scores. (**a**) Correlation of AI-based continuous scores with mean scores across three pathologists in the analytic performance test set. Results are shown for both AI-derived ordinal bins (blue) and pathologist-derived ordinal bins (gray). Filled circles indicate statistical significance, FDR-corrected *P* < 0.05. (**b**) cFib versus CPA measurements in primary endpoint responders in the ATLAS clinical trial. In primary endpoint responders, continuous fibrosis scores were significantly reduced in treated versus placebo patients (Mann–Whitney *U* = 20.0, *P* = 0.02), while proportionate area fibrosis measurements were not significantly reduced (Mann–Whitney *U* = 39.0, *P* = 0.21). (**c**) Stratification of patients with BL F3 or F4 fibrosis from STELLAR 3 and STELLAR 4 trial cohorts into rapid (red) and slow (orange) progressors based on continuous score cut-offs of 3.6 and 4.6, respectively. Kaplan–Meier and Cox proportional hazards regression analyses are shown. Rounded cutoffs were chosen to maximize hazards. (**d**) Discriminatory accuracy of AI-derived continuous scores versus ordinal scores to predict progression to cirrhosis (left) and LRE (right) in STELLAR 3 and STELLAR 4 trial cohorts. In both cases, using receiver operating characteristic analysis, the continuous AUC was significantly greater (progression to cirrhosis: 0.66 [95% CI: 0.60–0.71] versus 0.59 [95% CI: 0.55–0.60]; progression to LRE: 0.61 [95% CI: 0.51–0.71] versus 0.54 [95% CI: 0.47–59]). AI, artificial intelligence; AIM, artificial intelligence-based measurement; AUC, area under the receiver operating characteristic curve; BL, baseline; cFib, AI-derived continuous fibrosis scoring; CI, confidence interval; CPA, collagen proportionate area; CRN, Clinical Research Network; FDR, false discovery rate; FPR, false positive rate; LRE, liver-related event; NASH, nonalcoholic steatohepatitis; Tau, Kendall’s rank correlation coefficient for ordinal scores; TPR, true positive rate.

To further cross-validate the AI-derived continuous NASH CRN scores with other lines of clinical evidence, these continuous scores were correlated with the relevant corresponding noninvasive test (NIT) metrics in the ATLAS clinical trial dataset. NITs that were developed to serve as noninvasive biomarkers for specific histologic features or have previously been shown to correlate strongly with specific histologic features were correlated with the relevant continuous NASH CRN grades/stages (Supplementary Table 3). AI-derived continuous fibrosis stage was significantly correlated with liver stiffness by FibroScan (*r_τ_*: 0.33, *P* < 0.001), Fibrosis-4 (*r_τ_*: 0.23, *P* < 0.001), enhanced liver fibrosis (ELF) test (*r_τ_*: 0.22, *P* < 0.001), tissue inhibitor of metalloproteinases 1 (*r_τ_*: 0.11, *P* = 0.02), and amino terminal propeptide of type III procollagen (*r_τ_*: 0.14, *P* < 0.01), whereas continuous steatosis grade was not significantly correlated with the same NIT measures. Similarly, whereas continuous steatosis grade was significantly correlated with magnetic resonance imaging-proton density fat fraction (MRI-PDFF) (*r_τ_*: 0.52, *P* < 0.001), continuous fibrosis stage was not correlated with hepatic fat measured by MRI-PDFF (*r_τ_*: −0.11, *P* = 0.24). Continuous lobular inflammation grade was significantly correlated with C-reactive protein (*r_τ_*: 0.13, *P* < 0.01) and adiponectin levels (*r_τ_*: –0.15, *P* < 0.01), while continuous ballooning grade was significantly correlated with glycated hemoglobin (*r_τ_*: 0.16, *P* < 0.001). Notably, both continuous fibrosis stage and continuous steatosis grade were significantly correlated with collagen proportionate area (CPA) by morphometry, but in opposite directions (continuous fibrosis stage: *r_τ_*: 0.56, *P* < 0.001; continuous steatosis grade: *r_τ_*: −0.16, *P* < 0.001). This result is consistent with data showing a reduction in steatosis severity with progression of hepatic fibrosis in NASH^28–31^.

#### Advantage of AI-derived continuous NASH CRN fibrosis staging over standard continuous fibrosis measures

To assess whether AI-based continuous NASH CRN fibrosis staging captures greater changes in treatment versus placebo over conventional continuous fibrosis measures, we evaluated the relative sensitivity of the AI-based NASH CRN fibrosis staging model against an AI-derived proportionate area of fibrosis measurement (surrogate for CPA) in MT images from the ATLAS clinical trial dataset. The results showed that in primary endpoint responders, treated patients showed a significantly greater reduction in continuous fibrosis score (cFib) than placebo patients (Mann–Whitney *U* = 20.0, *P* = 0.02), whereas proportionate area fibrosis measurements were not significantly reduced in treated relative to placebo patients (Mann–Whitney *U* = 39.0, *P* = 0.21; Fig. 5b). In addition, cFib scores increased for many nonresponders but decreased for all responders, showing that the continuous scoring system was able to identify worsening fibrosis in patients not responding to treatment (Fig. 5b).

#### Advantage of continuous over ordinal scoring for predicting patient outcomes

To assess the potential utility of continuous NASH CRN scores compared with ordinal NASH CRN grades/stages for patient stratification and for predicting NASH patient outcomes, we examined the prognostic utility of continuous scoring for predicting progression to cirrhosis (F4) in patients with bridging (F3) fibrosis at baseline or predicting liver-related events (LREs) in patients with cirrhosis at baseline in the STELLAR 3 and STELLAR 4 NASH clinical trial cohorts^23^, respectively. Associations between continuous scores at baseline and clinical disease progression through end of follow-up were determined using the Kaplan–Meier method and Cox proportional hazards regression analysis, with rounded cutoffs selected to maximize hazards. We found that continuous fibrosis stage cutoffs of 3.6 and 4.6 maximized the stratification of patients into slow versus rapid progressors to cirrhosis or LREs, respectively (Fig. 5c). AI-derived continuous scoring showed higher discriminatory accuracy for predicting progression to cirrhosis and LREs than ML-derived ordinal scoring (Fig. 5d).

## Discussion

Pathologist assessment of liver histopathology is central to the evaluation of NASH disease severity and serves as the basis for patient selection and treatment efficacy assessment in NASH clinical trials. Histologic evaluation in NASH clinical trials has been limited by intra- and inter-pathologist variability in histologic grading and staging^6,32^. Both the FDA and NASH CRN have proposed panel scoring to reduce inter-pathologist variability; however, these guidelines have yet to be standardized or widely adopted, and variability in assessment, even among expert pathologists, remains high^6,7^.

We developed a suite of digital pathology algorithms that reproducibly predict the location, extent, and severity of histologic biomarkers of NASH via both AI-derived recapitulation of NASH CRN ordinal grading and staging, and novel AI-based qualitative and quantitative metrics. Our goal was to develop these tools to assist pathologists in locating and evaluating critical histologic signatures of NASH disease progression and regression. We also aimed to enable reproducible scoring that facilitates consistent measurement of changes in disease severity between baseline and end of treatment in NASH clinical trials.

We first demonstrated model performance accuracy using a clinical trial population by showing that AI-derived predictions for NASH CRN steatosis grade, lobular inflammation grade, ballooning grade, and fibrosis stage were concordant with expert pathologist consensus NASH CRN grading/staging in a NASH clinical trial population. AIM-NASH performance was tested by treating the model as an independent reader within a panel. Our results provide evidence that the model did not internalize any individual pathologist’s scoring biases, but instead learned to grade and stage histologic features in alignment with a consensus of expert NASH pathologists. These results suggest that AIM-NASH captured features and change in histology over time and in response to drug treatment in an unbiased manner that aligned with expert pathologist interpretation.

Furthermore, we demonstrated that model-derived ordinal scores recapitulated patient enrollment and endpoint measurement in a completed phase 2b NASH clinical trial. The model’s predictions were also highly reproducible when the analysis was repeated on the same images, suggesting that this approach could enable consistent measurement of disease severity within and across timepoints and clinical trials. Further analytical validation is being performed to assess reproducibility with various pre-analytic factors (different scanners, stain quality, biopsy and section quality). Together, the accuracy and reproducibility of AIM-NASH suggest that this AI-based system could be used in the future to assist pathologists in achieving reproducible grading/staging, especially for cases in which the disease pathology may reflect a boundary between two grades/stages, leading to discordance among pathologists.

We further tested the model’s utility as a research tool by performing a retrospective assessment of primary and exploratory study efficacy endpoints in the phase 2b ATLAS clinical trial^27^. We found that AIM-NASH, along with achieving a high level of scoring accuracy and superior repeatability compared with pathologists, detected a greater proportion of primary and exploratory endpoint responders in treated patients than manual scoring. Notably, this result was maintained when adjusting for the proportion of placebo patient responders. This finding was consistent with AIM-NASH-based retrospective analysis of other trial cohorts. We showed that AI, but not manual scoring, revealed statistically significant differences in response rates between treated and placebo patients in a completed phase 2 clinical trial, suggesting that the trial would have met its primary endpoint if histology had been assessed by AI^33^. Furthermore, we showed that placebo response rates were significantly reduced when assessed by AI relative to manual scoring in a phase 2 trial of patients with cirrhotic NASH^27,34^. Together, these results suggest that AIM-NASH CRN grading and staging of histologic features can detect histologic response to drug treatment with comparable accuracy to expert NASH pathologists, while providing greater sensitivity and reproducibility. AIM-NASH may thus have potential clinical utility for determining disease severity and may be applicable for more sensitive determination of drug efficacy in clinical trial settings. Future studies should include rigorous analytical validation to verify repeatability and reproducibility across scanners and scanner operators, in addition to clinical validation to verify the efficacy, utility, and scalability of an AI-assist clinical trial workflow.

Achieving the surrogate biopsy-based endpoints adopted by regulatory bodies for NASH clinical trials has been exceedingly difficult, owing to the slow rate at which NASH progresses and regresses^35^; consequently, no NASH therapeutic has been approved. To address this challenge, several measurement systems that detect subordinal levels of histologic change have been proposed^36^. We previously demonstrated the utility of AI-based continuous measures of fibrosis for detecting subtle, yet statistically significant changes in fibrosis in response to treatment^5,34^.

The continuous scoring system we present here maps each NASH CRN grade/stage to a bin derived from the standard NASH scoring system, allowing direct comparison between the ordinal and continuous scoring systems. The AI-derived continuous NASH CRN scores showed strong correlations with mean scores derived by a panel of expert NASH pathologists, where the directional bias of the panel was clearly reflected in the continuous score, and between these scores and relevant noninvasive NASH biomarkers that are known to correlate with specific histologic features and clinical outcomes^31^. The AIM-NASH–based continuous NASH CRN fibrosis score was more sensitive to treatment-induced changes in fibrosis than the gold standard continuous CPA and was more strongly predictive of patient progression to cirrhosis and liver-related complications than the AI-based ordinal NASH CRN grades/stages. Additionally, the continuous fibrosis score enabled the definition of cutoffs that stratified patients with NASH with stage 3 (F3) or stage 4 (F4) fibrosis into slow versus rapid disease progressors. These results suggest several important applications for continuous histologic scoring in NASH in both translational and clinical development settings.

One limitation of the continuous scoring system reported here is that it presents disease progression and regression on a linear scale; however, it is widely accepted that NASH disease neither progresses nor regresses linearly^35,37^. For instance, a change in continuous fibrosis stage from 3.0 to 3.2 may reflect a different amount of change in disease severity than a change from 4.0 to 4.2 or from 3.0 to 2.8. Future experiments should investigate whether this mapping of a nonlinear system to a linear scale complicates measurement of changes in disease severity in response to treatment, and whether it might be possible to develop a scale that more closely approximates the manner in which NASH disease progresses and regresses. Another limitation of the continuous scoring system is that clinically meaningful thresholds are not yet known. Additional research is needed to define and characterize meaningful changes in the continuous fibrosis score, such as a study to determine whether a sub-integer reduction in fibrosis score is associated with improved clinical outcome.

Importantly, the present results highlight the benefits of collaboration between AI developers and NASH pathologists to ensure that the technology being developed will improve pathologists’ evaluation of liver biopsies, while utilizing workflows that can be scaled to accommodate the increasing demand for NASH pathologists as the number of NASH clinical trials increases. The integration of AI-based digital pathology into NASH clinical trial workflows using validated WSI viewing platforms has the potential to facilitate more accurate identification of patients with NASH for trial enrollment, more robust measurement of histologic endpoints, and greater sensitivity to drug effect, which together promise to increase clinical trial success and improve patient outcomes.

## Data Availability

Codes for cell- and tissue-type model training, inference, and feature extractions are not disclosed. Access requests for such a code will not be considered to safeguard PathAI intellectual property. Access to histopathology features will be granted upon reasonable request from academic investigators without relevant conflicts of interest for non-commercial use who agree not to distribute the data. Access requests can be made to.

## Online methods

### Compliance

Artificial intelligence (AI)-based computational pathology models and platforms to support model functionality were developed using Good Clinical Practice/Good Clinical Laboratory Practice principles, including controlled process and testing documentation. All whole-slide images (WSIs) included in model development and performance evaluation are from patients who consented to the future use of these images for research purposes.

### Data collection

#### Datasets

Machine learning (ML) model development and external, held-out test sets are summarized in Supplementary Table 1. ML models for segmenting and grading/staging nonalcoholic steatohepatitis (NASH) histologic features were trained using 8747 haematoxylin and eosin (H&E) and 7660 Masson’s trichrome (MT) WSIs from six completed phase 2b and phase 3 NASH clinical trials, covering a range of drug classes, trial enrollment criteria, and patient statuses (screen fail versus enrolled) (Supplementary Table 1). H&E and MT liver biopsy WSIs from primary sclerosing cholangitis (PSC) and chronic hepatitis B infection were also included in model training. The latter dataset enabled the models to learn to distinguish between histologic features that may visually appear to be similar but are not as frequently present in NASH (e.g. interface hepatitis)^38^, in addition to enabling coverage of a wider range of disease severity than is typically enrolled in NASH clinical trials.

Model performance repeatability assessments and accuracy verification were conducted in an external, held-out validation dataset (Analytic performance test set) comprising WSIs of baseline and end-of-treatment (EOT) biopsies from a completed phase 2b NASH clinical trial (Supplementary Table 1). The clinical trial methodology and results have been described previously^22^. Digitized WSIs were reviewed for Clinical Research Network (CRN) grading and staging by the clinical trial’s three central pathologists (CPs), who have extensive experience evaluating NASH histology in pivotal phase 2 and 3 clinical trials and in the NASH CRN and European NASH pathology communities^6^. Images for which CP scores were not available were excluded from the model performance accuracy analysis. Median scores of the three pathologists were computed for all WSIs and used as a reference for AI model performance. Importantly, this dataset was not used for model development and thus served as a robust external validation dataset against which model performance could be fairly tested.

The clinical utility of model-derived features was assessed by generated ordinal and continuous ML features in WSIs from three completed NASH clinical trials: 1882 baseline and EOT WSIs from 395 patients enrolled in the ATLAS phase 2b clinical trial and 1519 baseline WSIs from patients enrolled in the STELLAR-3 (*n* = 725 patients) and STELLAR-4 (*n* = 794 patients) clinical trials. Dataset characteristics for these three trials have been published previously^27^.

#### Pathologists

Board-certified pathologists with a subspecialty in liver pathology assisted in the development of the present NASH AI algorithms by providing (a) hand-drawn annotations of key histologic features for training image segmentation models (see *Annotations*), (b) slide-level NASH CRN steatosis grades, ballooning grades, lobular inflammation grades, and fibrosis stages for training the AI scoring models (see *Model Development*), or (c) both. Pathologists who provided slide-level NASH CRN grades/stages for model development were required to pass a proficiency examination, in which they were asked to provide NASH CRN grades/stages for 20 NASH cases, and their scores were compared with a consensus median provided by three NASH CRN pathologists. Agreement statistics were reviewed by a PathAI expert NASH pathologist and leveraged to select pathologists for assisting in model development. In total, 59 pathologists provided feature annotations for model training; five pathologists provided slide-level NASH CRN grades/stages (see *Annotations*).

#### Annotations

*Tissue feature annotations*: Pathologists provided pixel-level annotations on WSIs using a proprietary digital WSI viewer interface. Pathologists were specifically instructed to draw, or “annotate,” over the H&E and MT WSIs to collect many examples of substances relevant to NASH, in addition to examples of artifact and background. Instructions provided to pathologists for select histologic substances are included in Supplementary Table 4. In total, 103,579 feature annotations were collected to train the ML models to detect and quantify features relevant to image/tissue artifact, foreground versus background separation, and NASH histology.

*Slide-level NASH CRN grading and staging*: All pathologists who provided slide-level NASH CRN grades/stages received and were asked to evaluate histologic features according to the Nonalcoholic Fatty Liver Disease Activity Score (NAS) and CRN Fibrosis Staging rubrics developed by Kleiner et al^16^. All cases were reviewed and scored using the aforementioned WSI viewer.

### Model development

#### Dataset splitting

The model development dataset described above was split into training (∼70%), validation (∼15%), and held-out test (∼15%) sets. The dataset was split at the patient level, with all WSIs from the same patient allocated to the same development set. Sets were also balanced for key NASH disease severity metrics, such as NASH CRN steatosis grade, ballooning grade, lobular inflammation grade, and fibrosis stage, to the greatest extent possible. The balancing step was occasionally challenging because of the NASH clinical trial enrollment criteria, which restricted the patient population to those fitting within specific ranges of the disease severity spectrum. The held-out test set contains a dataset from an independent clinical trial to ensure algorithm performance is meeting acceptance criteria on a completely held-out patient cohort in an independent clinical trial and avoiding any test data leakage^25^.

#### Convolutional neural networks

The present AI NASH algorithms were trained using the three categories of tissue compartment segmentation models described below. Summaries of each model and their respective objectives are included in Supplementary Table 5. For all convolutional neural networks (CNNs), cloud-computing infrastructure allowed massively parallel patch-wise inference to be efficiently and exhaustively performed on every tissue-containing region of a WSI, with a spatial precision of 4–8 pixels.

*Artifact Segmentation Model:* A CNN was trained to differentiate (1) evaluable liver tissue from WSI background and (2) evaluable tissue from artifacts introduced via tissue preparation (e.g. tissue folds) or slide scanning (e.g. out-of-focus regions). A single CNN for artifact/background detection and segmentation was developed for both H&E and MT stains (Fig. 1).

*H&E Segmentation Model*: For H&E WSIs, a CNN was trained to segment both the cardinal NASH H&E histologic features (macrovesicular steatosis, hepatocellular ballooning, lobular inflammation) and other relevant features, including portal inflammation, microvesicular steatosis, interface hepatitis, and normal hepatocytes (i.e. hepatocytes not exhibiting steatosis or ballooning; Fig. 1).

*MT Segmentation Models:* For MT WSIs, CNNs were trained to segment large intrahepatic septal and subcapsular regions (comprising non-pathologic fibrosis), pathologic fibrosis, bile ducts, and blood vessels (Fig. 1). All three segmentation models were trained utilizing an iterative model development process, schematized in Supplementary Fig. 2. First, the training set of WSIs was shared with a select team of expert liver pathologists who were instructed to annotate over the H&E and MT WSIs, as described above. This first set of annotations is referred to as “primary annotations.” Once collected, primary annotations were reviewed by internal pathologists, who removed annotations from pathologists who had misunderstood instructions or otherwise provided inappropriate annotations. The final subset of primary annotations was used to train the first iteration of all three segmentation models described above, and segmentation overlays (Fig. 2) were generated. Internal pathologists then reviewed the model-derived segmentation overlays, identifying areas of model failure and requesting correction annotations for substances for which the model was performing poorly. At this stage, the trained CNN models were also deployed on the validation set of images to quantitatively evaluate the model’s performance on collected annotations. After identifying areas for performance improvement, correction annotations were collected from expert pathologists to provide further improved examples of NASH histologic features to the model. Model training was monitored, and hyperparameters were adjusted based on the model’s performance on pathologist annotations from the held-out validation set until convergence was achieved and pathologists confirmed qualitatively that model performance was strong.

The artifact, H&E tissue, and MT tissue CNNs were trained using pathologist annotations comprising 8–12 blocks of compound layers with a topology inspired by residual networks and inception networks with a softmax loss^39–41^. Each CNN model’s learning was augmented using distributionally robust optimization^42,43^ to achieve model generalization across multiple clinical and research contexts and augmentations. Augmentations included, but were not limited to, vertical and horizontal flips, shifts in contrast, brightness, and saturation, and the introduction of artificial gaussian noise, blur, and sharpening. Input- and feature-level mix-up^44,45^ was also employed (as a regularization technique to further increase model robustness).

#### Graph neural networks

CNN model predictions were used in combination with NASH CRN scores from eight pathologists to train graph neural networks (GNNs) to predict ordinal NASH CRN grades for steatosis, lobular inflammation, ballooning, and fibrosis. GNN methodology was leveraged for the present development effort because it is well suited to data types that can be modeled by a graph structure, such as human tissues that are organized into structural topologies, including fibrosis architecture^46^. Here, the CNN predictions (WSI overlays) of relevant histologic features were clustered into “superpixels” to construct the nodes in the graph, reducing hundreds of thousands of pixel-level predictions into thousands of super-pixel clusters. WSI regions predicted as background or artifact were excluded during clustering. Directed edges were placed between each node and their five nearest neighboring nodes (via the K nearest neighbor algorithm). Each graph node was represented by three classes of features generated from previously trained CNN predictions predefined as biological classes of known clinical relevance. Spatial features included the mean and standard deviation of (x, y) coordinates. Topological features included area, perimeter, and convexity of the cluster. Logit-related features included the mean and standard deviation of logits for each of the classes of CNN-generated overlays. Scores from multiple pathologists were used for training, and consensus (*n* = 3) scores were used for evaluating model performance during training. Leveraging scores from multiple qualified NASH pathologists reduced the potential impact of scoring variability and bias associated with a single reader.

To further account for systemic bias, whereby some pathologists may consistently overestimate patient disease severity while others underestimate it, we specified the GNN model as a “mixed effects” model. Each pathologist’s policy was specified in this model by a set of bias parameters learned during training and discarded at test time. Briefly, to learn these biases, we trained the model on all unique label-graph pairs, where the label was represented by a score and a variable that indicated which pathologist in the training set generated this score. The model then selected the specified pathologist bias parameter and added it to the unbiased estimate of the patient’s disease state. During training, these biases were updated via backpropagation only on WSIs scored by the corresponding pathologists. When the GNNs were deployed, the labels were produced using only the unbiased estimate.

In contrast to our previous work, in which models were trained on scores from a single pathologist^5^, GNNs in this study were trained using NASH CRN scores from eight NASH pathologists on a subset of the data used for image segmentation model training (Supplementary Table 1). The GNN nodes and edges were built from CNN predictions of relevant histologic features in the first model training stage. This tiered approach improved upon our previous work, in which separate models were trained for slide-level scoring and histologic feature quantification. Here, ordinal scores were constructed directly from the CNN-labeled WSIs.

#### GNN-derived continuous score generation

Continuous NAS and CRN fibrosis scores were produced by mapping GNN-derived ordinal grades/stages to bins, such that ordinal scores were spread over a continuous range spanning a unit distance of 1 (Supplementary Fig. 2). Activation layer output logits were extracted from the GNN ordinal scoring model pipeline and averaged. The GNN learned inter-bin cutoffs during training, and piecewise linear mapping was performed per logit ordinal bin from the logits to binned continuous scores using the logit-valued cutoffs to separate bins. Bins on either end of the disease severity continuum per histologic feature have long-tailed distributions that are not penalized during training. To ensure balanced linear mapping of these outer bins, logit values in the first and last bins were restricted to minimum and maximum values, respectively, during a post-processing step. These values were defined by outer-edge cutoffs chosen to maximize the uniformity of logit value distributions across training data. GNN continuous feature training and ordinal mapping was performed for each NASH CRN NAS component fibrosis separately.

### Quality control measures

Several quality control (QC) measures were implemented to ensure model learning from high-quality data: (1) PathAI liver pathologists evaluated all annotators for annotation/scoring performance at project initiation; (2) PathAI pathologists performed QC review on all annotations collected throughout model training; following review, annotations deemed to be of high quality by PathAI pathologists were used for model training, while all other annotations were excluded from model development; (3) PathAI pathologists performed slide-level review of the model’s performance after every iteration of model training, providing specific qualitative feedback on areas of strength/weakness after each iteration; (4) model performance was characterized at the patch- and slide-levels in an internal (held-out) test set; (5) model performance was compared against pathologist consensus scoring in an entirely held-out test set, which contained images that were out of distribution relative to images from which the model had learned during development.

### Statistical analysis

#### Model performance repeatability

Repeatability of AI-based scoring (intra-method variability) was assessed by deploying the present AI algorithms on the same held-out analytic performance test set 10 times and computing percent positive agreement across the 10 reads by the model.

#### Model performance accuracy

To verify model performance accuracy, model-derived predictions for ordinal NASH CRN steatosis grade, ballooning grade, lobular inflammation grade, and fibrosis stage were compared with median consensus grades/stages provided by a panel of three expert NASH pathologists who had evaluated NASH biopsies in a recently completed phase 2b NASH clinical trial (Supplementary Table 1). Importantly, images from this clinical trial were not included in model training and served as an external, held-out test set for model performance evaluation. Alignment between model predictions and pathologist consensus was measured via agreement rates, reflecting the proportion of positive agreements between the model and consensus.

We also evaluated the performance of each expert reader against a consensus to provide a benchmark for algorithm performance. For this mixed leave-one-out (MLOO) analysis, the model was considered a fourth “reader,” and a consensus, determined from the model-derived score and that of two pathologists, was used to evaluate the performance of the third pathologist left out of the consensus. The average individual pathologist versus consensus agreement rate was computed per histologic feature as a reference for model versus consensus per feature. Confidence intervals were computed using bootstrapping. Concordance was assessed for scoring of steatosis, lobular inflammation, hepatocellular ballooning, and fibrosis using the NASH CRN system.

#### AI-based assessment of clinical trial enrollment criteria and endpoints

The analytic performance test set (Supplementary Table 1) was leveraged to assess the AI’s ability to recapitulate NASH clinical trial enrollment criteria and efficacy endpoints. Baseline and EOT biopsies across treatment arms were grouped, and efficacy endpoints were computed using each study patient’s paired baseline and EOT biopsies. For all endpoints, the statistical method used to compare treatment with placebo was a Cochran–Mantel–Haenszel test, and *P* values were based on response stratified by diabetes status and cirrhosis at baseline (by manual assessment). Concordance was assessed with k statistics, and accuracy was evaluated by computing F1 scores. A consensus determination (*n* = 3 expert pathologists) of enrollment criteria and efficacy served as reference for evaluating AI concordance and accuracy. To evaluate the concordance and accuracy of each of the three pathologists, AI was treated as an independent, fourth “reader”, and consensus determinations were composed of AIM and two pathologists for evaluating the third pathologist not included in the consensus. This MLOO approach was followed to evaluate the performance of each pathologist against a consensus determination.

#### Continuous score interpretability

To demonstrate interpretability of the continuous scoring system, we first generated NASH CRN continuous scores in WSIs from a completed phase 2b NASH clinical trial (Supplementary Table 1, Analytic performance test set). We then compared the continuous scores across all four histologic features with the mean pathologist scores from the three study central readers, using Kendall rank correlation. The goal in measuring the mean pathologist score was to capture the directional bias of this panel per feature and verify whether the AI-derived continuous score reflected the same directional bias.

## Acknowledgments

Medical writing support and editorial assistance were provided by Sandra J Page, PhD, and Agata Shodeke, PhD, of Spark Medica Inc, according to Good Publication Practice guidelines, funded by PathAI.

---

**Supplementary Table 1.** Model development and test dataset characteristics. ASK1, apoptosis signal-regulating kinase 1 (also known as mitogen-activated protein kinase kinase kinase 5); F, fibrosis stage; HBV, hepatitis B virus; H&E, hematoxylin and eosin; LOXL2, lysyl oxidase-like 2; MT, Masson’s trichrome; NAS, NAFLD activity score; NASH, nonalcoholic steatohepatitis; PPARδ, peroxisome proliferator activated receptor delta; PSC, primary sclerosing cholangitis; WSI, whole slide image.

| Dataset | Trial phase | Number of WSIs (H&E, MT) | Drug class | Enrollment criteria |
| --- | --- | --- | --- | --- |
| Model development datasets |  |  |  |  |
| 1 | 3 | 2188, 2188 | ASK1 inhibitor | NASH diagnosis; fibrosis F3<sup>23</sup> |
| 2 | 3 | 2488, 2478 | ASK1 inhibitor | NASH diagnosis; fibrosis F4<sup>23</sup> |
| 3 | 2b | 528, 528 | Monoclonal antibody directed against LOXL2 | NASH defined as steatosis > 5% with associated lobular inflammation: Ishak stage 3,4<sup>47</sup> |
| 4 | 2b | 561, 554 | Monoclonal antibody directed against LOXL2 | NASH diagnosis; Ishak stage 5,6<sup>21</sup> |
| 5 | 2 | 158, 163 | ASK1 Inhibitor, monoclonal antibody directed against LOXL2 | Evidence of NASH with fibrosis on biopsy<sup>48</sup> |
| 6 | 2 | 312, 312 | PPARδ agonist | Definite NASH; NAS ≥ 4 with 1 per component; fibrosis F1, F2, F3<sup>49</sup> |
| 7 | 3 | 1477, 766 | Nucleotide analogue (antiviral) | HBV<sup>50</sup> |
| 8 | 3 | 851, 415 | Nucleotide analogue (antiviral) | HBV<sup>50</sup> |
| 9 | 2b | 331, 333 | Monoclonal antibody directed against LOXL2 | PSC<sup>24</sup> |
| Analytic performance test set |  |  |  |  |
| 10 | 2b | 639, 633 | Insulin sensitizer | Definite NASH; NAS ≥ 4 with 1 per component; Fibrosis F1, F2, F3<sup>22</sup> |

ASK1, apoptosis signal-regulating kinase 1 (also known as mitogen-activated protein kinase kinase kinase 5); F, fibrosis stage; HBV, hepatitis B virus; H&E, hematoxylin and eosin; LOXL2, lysyl oxidase-like 2; MT, Masson's trichrome; NAS, NAFLD activity score; NASH, nonalcoholic steatohepatitis; PPARδ , peroxisome proliferator activated receptor delta; PSC, primary sclerosing cholangitis; WSI, whole slide image.

**Supplementary Table 2.** Algorithm repeatability assessment using 10 independent reads per WSI. WSI, whole slide image.

|  | Number of WSIs | Model versus model agreement rate |
| --- | --- | --- |
| Steatosis | 639 | 100% |
| Lobular inflammation | 639 | 100% |
| Ballooning | 639 | 100% |
| Fibrosis | 633 | 100% |

WSI, whole slide image.

**Supplementary Table 3.** Correlations between the AI-derived continuous scoring system and comparable noninvasive tests. AI, artificial intelligence; ELF, enhanced liver fibrosis test; FIB4, fibrosis-4; HbA1C, hemoglobin A1c; MRI-PDFF, magnetic resonance imaging derived proton density fat fraction; NIT, noninvasive test; PIIINP, procollagen III N-terminal peptide; Kendall’s Tau, Kendall’s rank correlation coefficient for ordinal scores; TIMP, tissue inhibitor of metalloproteinase; TIMP1, TIMP metallopeptidase inhibitor 1.

| Continuous scoring system | NIT | Kendall's Tau | P value | n |
| --- | --- | --- | --- | --- |
| Continuous fibrosis stage | FibroScan | 0.33 | 2.49E−11 | 188 |
| Continuous fibrosis stage | FIB4 | 0.23 | 1.56E−06 | 207 |
| Continuous fibrosis stage | ELF | 0.22 | 2.52E−06 | 210 |
| Continuous fibrosis stage | TIMP1 | 0.11 | 2.01E−02 | 210 |
| Continuous fibrosis stage | PIIINP | 0.14 | 3.03E−03 | 210 |
| Continuous fibrosis stage | MRI-PDFF | −0.11 | 2.36E−01 | 59 |
| Continuous fibrosis stage | Morphometric quantitative collagen (%) | 0.56 | 2.20E−32 | 205 |
| Continuous steatosis grade | MRI-PDFF | 0.52 | 4.83E−09 | 59 |
| Continuous steatosis grade | Morphometric quantitative collagen (%) | −0.16 | 5.42E−04 | 205 |
| Continuous lobular inflammation grade | C-reactive protein | 0.13 | 5.04E−03 | 211 |
| Continuous lobular inflammation grade | Adiponectin | −0.15 | 1.38E−03 | 211 |
| Continuous ballooning grade | HbA1C | 0.16 | 8.36E−04 | 211 |

AI, artificial intelligence; ELF, enhanced liver fibrosis test; FIB4, fibrosis-4; HbA1C, hemoglobin A1c; MRI-PDFF, magnetic resonance imaging derived proton density fat fraction; NIT, noninvasive test; PIIINP, procollagen III N-terminal peptide; Kendall's Tau, Kendall's rank correlation coefficient for ordinal scores; TIMP, tissue inhibitor of metalloproteinase; TIMP1, TIMP metalloproteinase inhibitor 1.

**Supplementary Table 4.** Example instructions for the interpretation of histologic features.

| Histologic feature | Example instructions |
| --- | --- |
| Lobular inflammation | Place label regions containing at least three inflammatory cells, not including those within sinusoids. Do not label regions of portal inflammation with this region label. |
| Hepatocyte ballooning | Please use this label on regions of hepatocellular ballooning. Hepatocellular ballooning is defined as round cells with rarified cytoplasm that are at least 50% larger than neighboring normal cells. |
| Steatosis | Please use this label on regions of dense steatosis. |
| Thick pathologic fibrotic septa | Please use this label for thickened fibrotic septae extending from portal and central regions considered when staging liver biopsies. |
| Portal tract (normal) | Please use this label for normal-appearing, small-/medium-sized portal regions, not expanded by fibrosis or inflammation. |
| Portal tract (abnormal) | Please use this label for portal regions expanded by inflammation, fibrosis, bile ductular proliferation, or any combination of the above. |
| Large normal septa | Please use this label for larger intrahepatic normal septae (usually containing larger arteries, veins, and bile ducts) that would not be included when staging liver biopsies. |
| Subcapsular fibrosis | Please use this label for normal subcapsular regions of fibrosis not considered when staging liver biopsies. |

**Supplementary Table 5.**
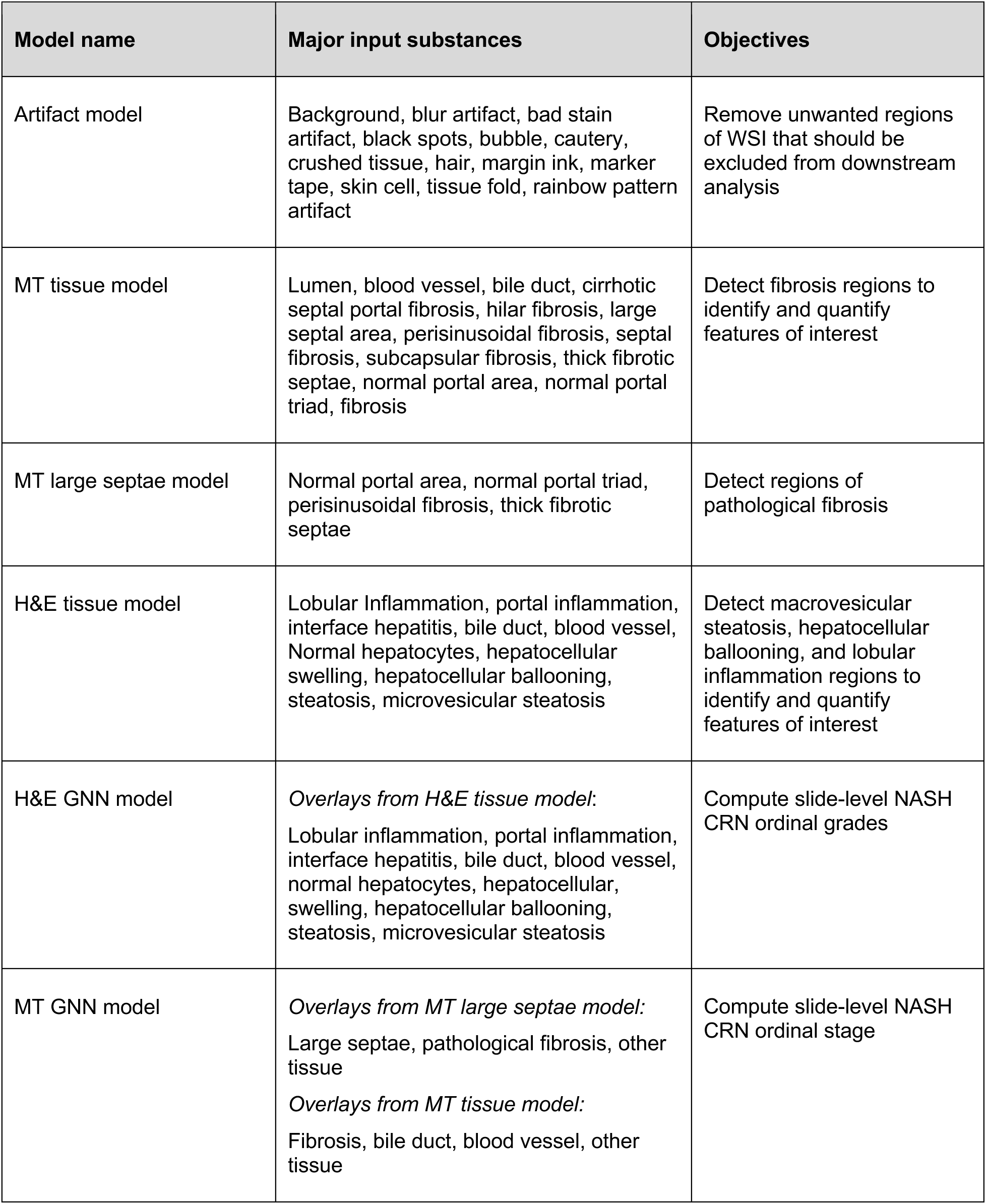
AI-derived models, input substances, and objectives for application. AI, artificial intelligence; CRN, Clinical Research Network; GNN, graph neural network; H&E, hematoxylin and eosin; MT, Masson’s trichrome; NAS, nonalcoholic fatty liver disease activity score; NASH, nonalcoholic steatohepatitis; WSI, whole slide image.

**Supplementary Fig. 1.**
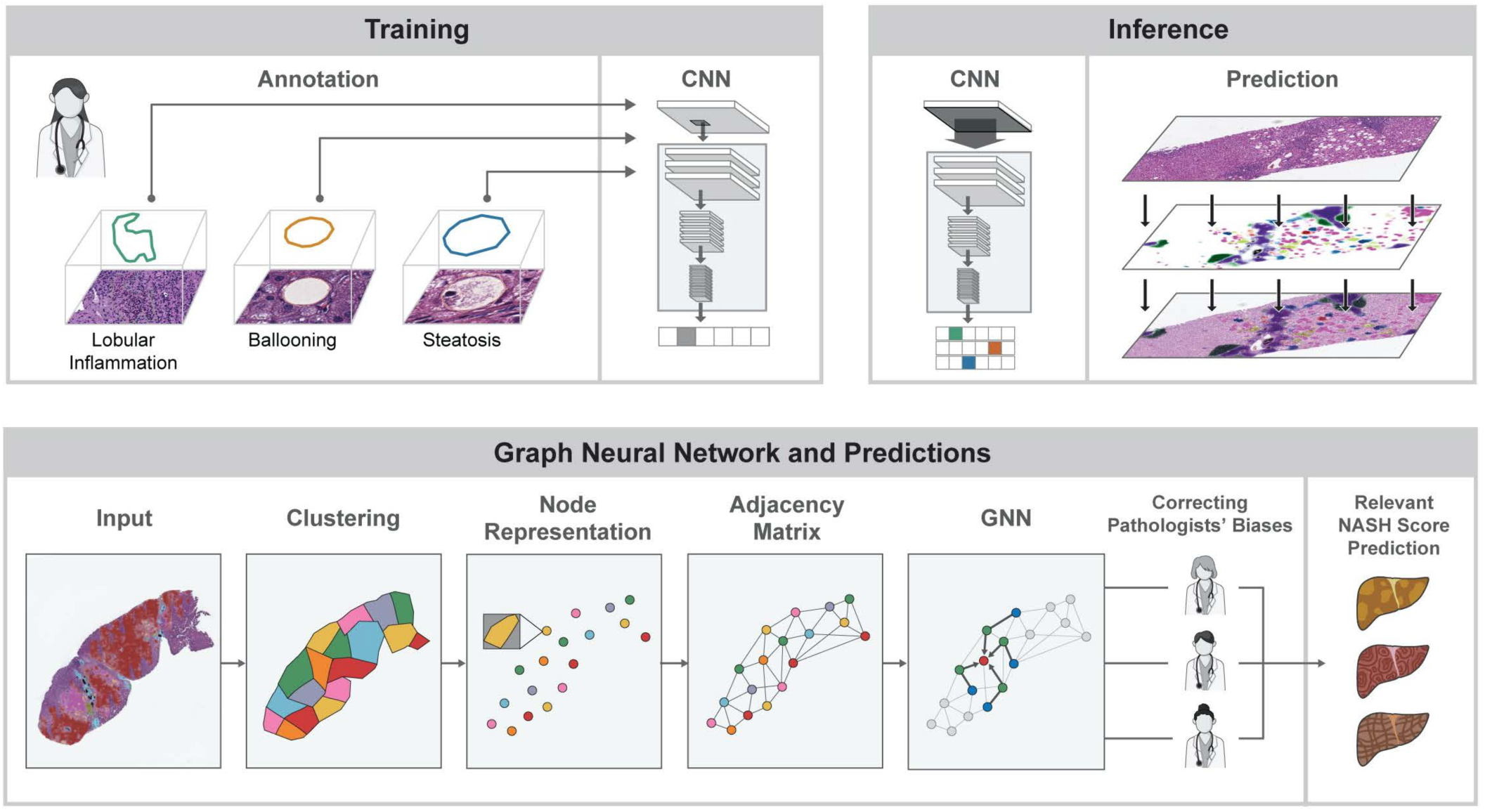
Model training schematic. CNN, convolutional neural network; GNN, graph neural network; NASH, nonalcoholic steatohepatitis.

**Supplementary Fig. 2.**
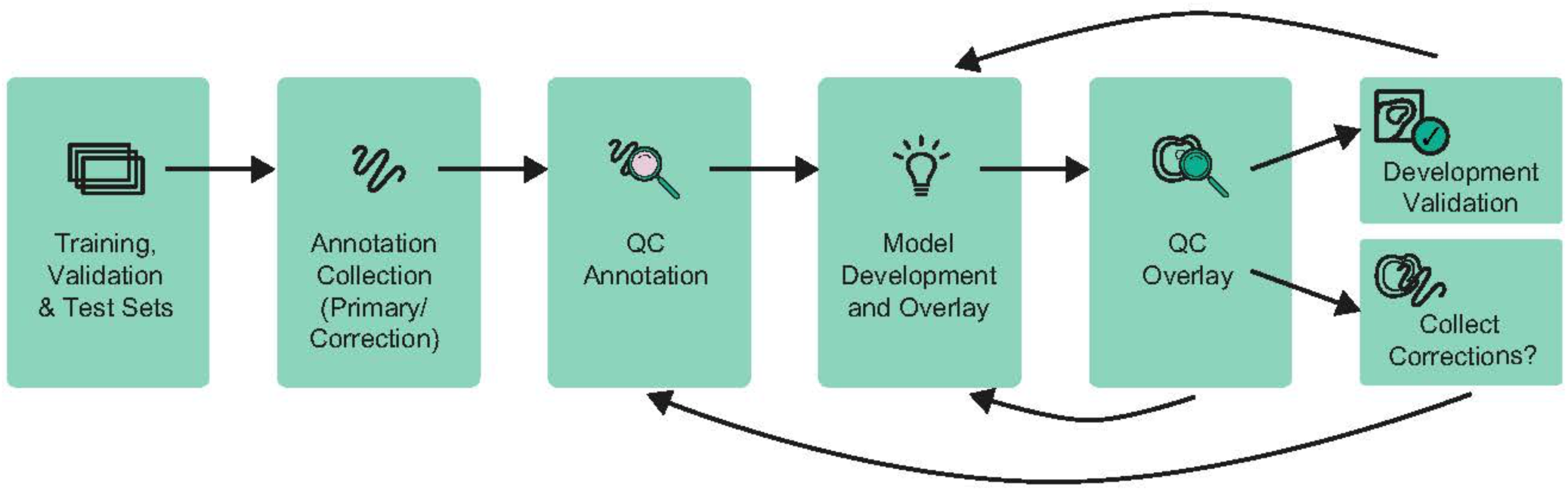
Segmentation model development process. QC, quality control.

**Supplementary Fig. 3.**
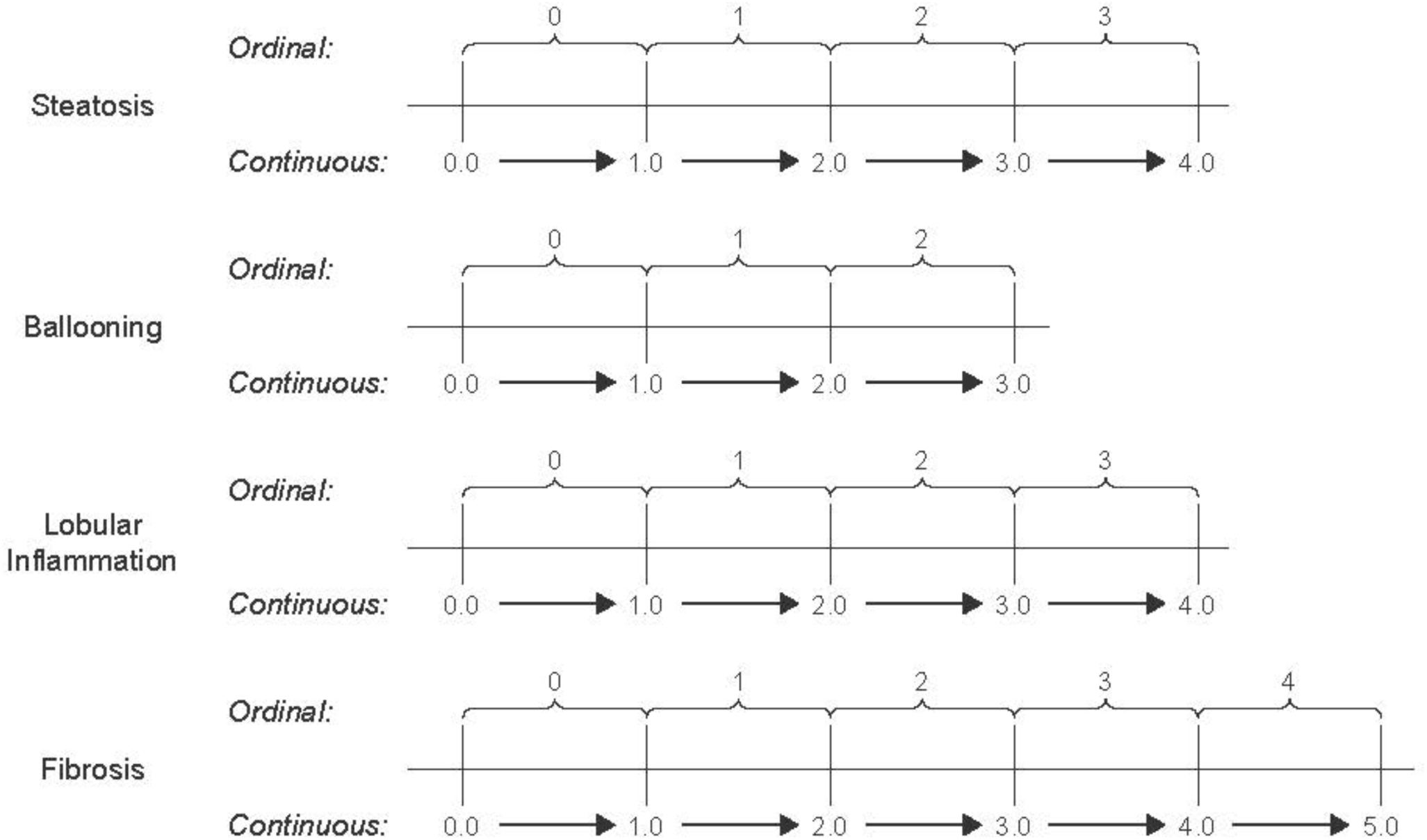
Mapping of continuous scores.

